# Alzheimer’s disease clinical decision points for two plasma p-tau217 laboratory developed tests in neuropathology confirmed samples

**DOI:** 10.1101/2024.07.27.24310872

**Authors:** Anna E. Mammel, Ging-Yuek Robin Hsiung, Ali Mousavi, Kelsey Hallett, Ian R. Mackenzie, Veronica Hirsch-Reinshagen, Don Biehl, Pradip Gill, Mary Encarnacion, Hans Frykman

## Abstract

**INTRODUCTION:** We evaluated the diagnostic performance of two commercial plasma p-tau217 immunoassays compared to CSF testing and neuropathology.

**METHODS:** 170 plasma samples from University of British Columbia Hospital Clinic for Alzheimer’s (AD) and Related Disorders were analyzed for p-tau217 using Fujirebio and ALZpath assays. Decision points were determined using CSF testing and autopsy findings as the standard.

**RESULTS:** Fujirebio and ALZpath p-tau217 had similar overall analytical and clinical performance, with distinct decision points for each assay. Based on autopsy finding, both p-tau217 assays identified individuals with AD from other neurodegenerative diseases (ALZpath AUC = 0.94, Fujirebio AUC= 0.90). The ALZpath assay detected AD pathology at milder disease stages compared to the Fujirebio assay.

**DISCUSSION:** Our study reinforces the clinical utility of plasma p-tau217 as an AD biomarker. Differences in test performance and clinical decision points suggest an assay specific diagnostic approach is required for plasma p-tau217 in clinical practice.

## 1. BACKGROUND

Alzheimer disease (AD) is characterized biologically by Aβ plaques and tau neurofibrillary tangles. ^1,2^ AD pathology can frequently co-occur with other neurodegenerative and vascular diseases, and accurate timely differential diagnosis is critical for appropriate care and precise treatment strategies. ^1,3–6^ In recent years pathophysiologic and topographical biomarkers have significantly improved diagnosis, clinical evaluation, and disease monitoring of AD patients. ^7^ Among pathophysiologic biomarkers, amyloid PET, fluid biomarkers such as CSF concentrations of Aβ and tau proteins, along with plasma concentrations of Aβ and tau, all play a crucial role in this diagnostic enhancement.^7^

While invasiveness, accessibility, cost, and availability limit the use of PET and CSF biomarkers in clinical practices, the development of novel immunoassay technologies has facilitated the early detection of biomarkers such as phosphorylated tau in plasma. A number of very sensitive p-tau assays for measuring AD blood biomarkers, including p-tau181, p-tau217, and p-tau231, have recently been developed. ^8^All available p-tau assays measure phosphorylated forms of tau using specific antibodies that target the N-terminus or mid-domain of the protein with higher concentrations than the full-length or C-terminal of tau in biofluids.^9^

Several large clinical-based studies also have shown that plasma p-tau217 can distinguish AD from other neurodegenerative diseases with high diagnostic accuracy. ^10^P-tau217 can differentiate AD cases with significant brain pathology from non-AD cases, and other tauopathies. ^11^Furthermore, plasma p-tau217 is correlated more firmly with brain β-amyloid and paired helical filament tau tangles than plasma p-tau181.^12^ Several studies suggested that plasma p-tau217 is an accurate marker to predict the future development of AD in symptomatic patients with either MCI or subjective cognitive decline, and it is better for AD diagnosis relative to its large increase in the earlier stage of AD.^13–15^ ^16,17^

With the recent advancement in new AD-modifying treatments and the role of plasma p-tau217 in Alzheimer’s diagnosis, it is crucial to establish a reliable immunoassay for measuring plasma p-tau217 in the clinical laboratory. Several assays such as the Janssen SIMOA immunoassay using the Quanterix HD-X platform, and the Lilly immunoassay using the MSD ECL platform, have been developed for measuring plasma p-tau217. ^18,19^ However, these are “homebrewed” using proprietary antibodies and have not been made widely available for clinical testing since their initial development for research. More recently, the ALZpath p-tau217 assay, which is a SIMOA immunoassay for use on the Quanterix HD-X platform, has been made commercially available as a research use only (RUO) assay. A recently published paper highlighted how the efficacy of the plasma ALZpath p-tau217 immunoassay in accurate diagnosis of biological AD was similar to CSF biomarkers and PET. ^20^This study identified reproducible cut-offs across three cohorts to detect longitudinal alterations of plasma p-tau217, particularly at the preclinical stage. However, the clinical utilization of ALZpath p-tau217 immunoassay by SIMOA, as a laboratory diagnostic test (LDT), required additional validation studies, which are described in this study.

Additionally, the Fujirebio Lumipulse G instrument is a fully automated platform that has been used to measure CSF AD biomarkers in clinical laboratories. Fujirebio recently launched assays to measure p-tau217 in plasma.^21^ This assay is currently being used by national clinical laboratories to measure plasma levels of p-tau217 in a clinical setting as an LDT without FDA approval. Given the numerous studies demonstrating the reliability of plasma p-tau217 to distinguish AD pathology from non-AD conditions, our study focuses on two commercially available laboratory developed tests for plasma p-tau217, ALZpath p-tau 217 and Fujirebio p-tau217. Furthering the work of a recent head-to-head studies evaluating the diagnostic accuracy of ALZpath and Fujirebio plasma p-tau217 assays in clinically diagnosed AD^22,23^, we assessed the diagnostic performance of both assays in comparison to FDA-approved CSF testing for amyloid Aß42/40 ratio and p-tau181, as well as postmortem neuropathological evaluation of cases who had paired plasma samples at diagnosis of dementia.

## 2. METHODS

### 2.1 Study population

Participants were recruited from the University of British Columbia hospital clinic for Alzheimer and related disorders (UBCH-CARD) and at St. Paul’s Hospital, in Vancouver, Canada. Patients are assessed by specialty-trained neurologists and geriatricians who have expertise in dementia assessments. After initial detailed evaluation, patients are clinically diagnosed as AD, based on NIA-AA criteria or other established clinical diagnoses criteria. ^24^

### 2.2 Plasma and CSF sampling

Venous blood and CSF collection occurred on the same day within 4 hours. Specimens were collected by a standardized protocol based on the ADNI biobanking procedures (https://adni.loni.usc.edu/methods/). Venous blood samples were collected with K2 EDTA collection tube (BD vacutainers ® purple top), which were centrifuged at 2500 g for 15 minutes at 4^0^C, and the plasma supernatant was aliquoted into polypropylene tubes (200 µL tubes, Sarstedt AG & Co.) and stored at −80 C and thawed only once at the time of analysis. All CSF and plasma samples were evaluated in a single laboratory (Neurocode USA Inc., USA) on either the Lumipulse G1200 or HD-X Analyzer platform. Plasma p-tau217 was measured using two commercially available immunoassays, ALZpath Simoa p-tau217 v2 kits (Quanterix, MA USA) on the Quanterix HD-X Analyzer platform and Lumipulse G p-tau217 Plasma kits (Fujirebio Europe N.V., Belgium) on the Lumipulse G1200 platform.

CSF was collected in low-bind 15 mL tubes (Falcon® Conical Centrifuge polypropylene tubes, Corning Life Sciences), and after centrifugation at 4^0^C, the CSF was aliquoted into 200 microL polypropylene tubes (200 microL tubes, Sarstedt AG & Co.) and stored at −80 C until analysis. CSF Aβ 42 to 40 ratio and p-tau181 levels were measured using commercially available Lumipulse G β-Amyloid 1-40, Lumipulse G β-Amyloid 1-42, and Lumipulse G p-tau181 kits (Fujirebio Europe N.V., Belgium) on the Lumipulse G1200 platform. Performance assessment of β-Amyloid 1-40, β-Amyloid 1-42, and p-tau181 was verified by Neurocode USA, Inc.

### 2.3 Analytical validation of the p-tau217 assays

The analytical validation of both p-tau217 immunoassays was conducted by Neurocode at their CLIA licensed facility in Bellingham, WA USA. The sensitivity (limit of blank, limit of detection, and lower limit of quantification), precision, linearity, recovery, interference, and sample stability were validated according to methods developed by the Clinical and Laboratory Standards Institute (CLSI). Limit of blank, detection, and quantification were determined based on CLSI EP17-A2E guidelines. Limit of blank was calculated for each assay using the 95^th^ percentile of at least 60 blank samples. Limit of detection was determined for the ALZpath and Fujirebio p-tau217 assay using a parametric approach. For both the Fujirebio and ALZpath p-tau217 assays, the lower limit of quantification was determined by creating a precision profile using 4 low-level healthy control samples and the 4 precision study samples.

Precision was determined based on CLSI EP05-A3 guidelines. Precision was calculated utilizing data from over at least 20 runs, using internal quality control (IQC) samples made from a pool of three to four EDTA plasma samples. Dilution linearity was assessed using CLSI EP06-A guidelines, by serially diluting a high pooled sample into a healthy control sample and comparing the expected values with the observed values. Recovery was assessed using three samples spiked with an assay-specific spike (either calibrator concentrate or a manufacturer-provided control sample) that were measured diluted both with the spike and without and calculating percent recovery using CLSI EP34. Interference was assessed by spiking healthy control samples with maximum concentrations of interferents: unconjugated and conjugated bilirubin (Millipore), hemoglobin (Sigma-Aldrich), intralipid (Sigma-Aldrich), and heterophilic antibodies (Roche) per CLSI EP07-A3 guidelines. Sample stability was determined for each assay per CLSI EP25-A guidelines, by measuring samples at room temperature, 4 C, −20 C, and −80 C for various timepoints, and calculating the percent difference from the baseline concentration. Samples were also tested for freeze-thaw stability up to 5 times, calculating the percent difference from the baseline concentration.

### 2.4 Neuropathology evaluation

We examined 115 cases that had autopsies performed, allowing pathological confirmation of their clinical diagnoses. The autopsies were performed at the Department of Pathology and Laboratory Medicine on patients who were referred from the University of British Columbia Hospital Clinic for Alzheimer’s and Related Disorders (UBCH-CARD) with informed consent. Formalin-fixed, paraffin-embedded brain tissue blocks were cut at 5 microns and stained with hematoxylin and eosin (HE), HE combined with Luxol fast blue, modified Bielschowsky silver, Gallyas silver, and Congo red stains. Standard immunohistochemistry was performed using the Dako Omnis automated staining system with primary antibodies against alpha-synuclein (α-syn) (Thermo Scientific; 1:10,000 following microwave antigen retrieval), beta-amyloid (DAKO; 5 1:100 with initial incubation for 3 h at room temperature), hyperphosphorylated tau (clone AT-8; Innogenetics, Ghent, Belgium; 1:2000 following microwave antigen retrieval), phosphorylation independent TDP-43 (ProteinTech; 1:1000 following microwave antigen retrieval), and ubiquitin (DAKO; 1:500 following microwave antigen retrieval). The severity of senile neuritic plaque pathology was staged according to Consortium to Establish a Registry for Alzheimer’s Disease (CERAD) recommendations. The extent of tau pathology was staged according to Braak staging.^25–29^

### 2.5 Statistical analysis

Statistical analysis was performed in R (version 4.3.1 <u>R: The R Project for Statistical Computing</u> (r-project.org)). Accuracy and clinical decision point analysis was performed using receiver operating characteristic (ROC) curves generated using the R package pROC (<u>CRAN - Package pROC</u> (r-project.org)). Delong’s test for two correlated ROC curves was used to compared test performance. Linear correlation was determined using Spearman correlation test. Comparison of two sets of variables was conducted using a Welch’s two sample t-test. For three or more normally distributed continuous variables, ANOVA was used followed by a pair-wise comparison using Tukey’s HSD test.

## 3. RESULTS

### 3.1 Analytical performance of two p-tau217 plasma immune assays

ALZpath and Fujirebio p-tau217 assays were validated as laboratory developed tests based on CLSI guidelines at Neurocode laboratory. All plasma samples measured for p-tau217 by the ALZpath assay were above the lower limit of quantification (0.032 pg/mL). For p-tau217 plasma measured by the Fujirebio assay, 10 samples (6.5%) had values below the limit of quantification (0.06 pg/mL). The plasma concentrations ranged from 0.041 to 3.19 pg/mL for the Fujirebio assay and 0.11 to 3.50 pg/mL for the ALZpath assay. Intra-laboratory coefficient of variation was assessed for each assay using three concentration levels (low, medium, and high) and were 12.1%, 12.2%, and 5.3%, respectively (N = 35 per sample) for Fujirebio. For ALZpath the coefficient of variation for a low, medium, and high sample was 10.4%, 10.4%, and 9.9%, respectively (N = 53 per sample) (Table 1). Sample stability and interference was similar between the two plasma p-tau217 assays, though moderate heterophilic antibodies interference and reduced frozen sample stability at −20 C was observed for the Fujirebio assay (Table 1). Overall, the analytical performance was found to be comparable between the two p-tau217 assays (Table 1).

**Table 1:** Plasma p-tau217 immunoassay analytical validation summary.

| Analytical performance | ALZpath p-tau217 | Lumipulse p-tau217 |
| --- | --- | --- |
| Limit of blank (LoB) | 0.0035 ng/L | 0.040 ng/L |
| Limit of detection (LoD) | 0.0074 ng/L | 0.052 ng/L |
| Limit of quantification (LoQ) | 0.032 ng/L | 0.060 ng/L |
| Clinical reportable range | 0.032 – 10.00 ng/L | 0.06 – 10.00 ng/L |
| Intra-laboratory precision | ≤ 10.4% CV above LLoQ | ≤ 12.2% CV above LLoQ |
| Sample Stability | ≤ 7 days at 2 - 8°C<br>≤ 4 weeks -20°C<br>≤ 5 freeze/thaw cycles | ≤ 7 days at 2 - 8°C<br>≤ 1 weeks -20°C<br>≤ 3 freeze/thaw cycles |
| Interference | No interference detected to the maximum concentration for bilirubin (unconjugated and conjugated), hemoglobin, intralipid, biotin, and heterophilic antibodies | No interference detected to the maximum concentrations tested for bilirubin (unconjugated and conjugated), hemoglobin, and intralipid. Interference was detected for heterophilic antibodies |

### 3.2 Study participant clinical characteristics

A total of 170 participants were included in this study. All participants had plasma samples collected, 55 had corresponding CSF samples, and a distinct cohort of 115 had brain autopsy examination by a neuropathologist. The CSF subgroup had the following demographics: mean age 67.2 ± 10.4 years, 49.1% female, 47.7% APOE ε4 carriers, as described in Table 2. Amyloid status was determined by CSF Aβ42/40, 35 subjects (63.6%) were amyloid positive. Tau status was determined by CSF p-tau181, 31 subjects (56.4%) were tau positive. Five subjects were positive for amyloid only and one subject was positive for tau only. Amyloid and tau negative subjects had a mean CSF Aβ 42/40 ratio of 0.092 ± 0.010 and CSF p-tau181 concentration of 28.4 ± 10.0 pg/mL. Amyloid and tau positive subjects had a mean CSF Aβ 42/40 ratio of 0.045 ± 0.010 and a mean CSF p-tau181 concentration of 90.0 ± 28.7 pg/mL (Table 3).

**Table 2:** Characteristics of the CARD neuropathology (NP) and CSF cohorts.

| Characteristic | Mean (SD) |  |
| --- | --- | --- |
|  | NP CARD<br>(N = 115) | CSF CARD<br>(N = 55) |
| Age, y* | 71.8 (11.9) | 67.2 (10.4) |
| Sex, no. (%) |  |  |
| Female | 54 (47.0) | 27 (49.1) |
| Male | 61 (53.0) | 28 (50.9) |
| APOE ε4 carriers, no. (%) | 57 (49.6) | 21 (47.7) † |
| Diagnosis, no. (%)‡ |  |  |
| AD only | 30 (26.1) | 21 (38.2) |
| AD with CAA | 7 (6.1) | -- |
| AD with DLB | 13 (11.3) | 1 (1.8) |
| AD with Vascular | 7 (6.1) | -- |
| AD with mix pathologies | 13 (11.3) | 4 (7.2) |
| MCI | -- | 13 (23.6) |
| NCI | -- | 2 (3.6) |
| TDP-43 only | 18 (15.7) | -- |
| Synuclein only | 4 (3.5) | -- |
| Tau only | 18 (15.7) | 1 (1.8) |
| other non-AD | 5 (4.3) | 13 (3.6) |
| CSF Aβ 42/40, pg/mL | -- | 0.063 (0.025) |
| CSF p-tau181, pg/mL | -- | 63.4 (37.4) |
| Plasma p-tau217 (ALZpath), pg/mL | 0.91 (0.74) | 0.83 (0.76) |
| Plasma p-tau217 (Lumipulse), pg/mL | 0.54 (0.58) § | 0.44 (0.49) ¶ |

\* Age refers to age at death for the neuropathy cohort and age at fluid biomarker collection for CSF cohort.
† N = 44 samples with APOE genotyping
‡ Diagnosis refers to final diagnosis for the neuropathy cohort and clinical diagnosis for the CSF cohort.
§ Insufficient volume for testing in a subset of plasma samples, N = 102.
¶ Insufficient volume for testing in a subset of plasma samples, N = 51.

**Table 3:** Fluid biomarker concentrations by amyloid and tau status.

| AT | N (%) | CSF mean ± SD pg/mL | | Plasma p-tau217 mean ± SD pg/mL | |
| --- | --- | --- | --- | --- | --- |
| | | Aβ 42/40 | p-tau181 | ALZpath | Fujirebio |
| A-T- | 19 (34.5%) | 0.092 ± 0.01 | 28.4 ± 10.0 | 0.26 ± 0.13 | 0.10 ± 0.09 |
| A-T+ | 1 (1.8%) | 0.091 | 60.5 | 0.33 | 0.14 |
| A+T- | 5 (9.1%) | 0.059 ± 0.02 | 37.1 ± 9.39 | 0.50 ± 0.28 | 0.18 ± 0.11 |
| A+T+ | 30 (54.5%) | 0.045 ± 0.01 | 90.0 ± 28.7 | 1.29 ± 0.79 | 0.72 ± 0.53 |

The neuropathology subgroup had a mean age at death of 71.8 ± 11.9 years, 47.0% of the subjects were female, and 49.9% were APOE e4 carriers. Final diagnosis was a mixture of pure AD (N = 30 (26.1%)), AD with mixed pathology (N = 42 (36.5%)), and non-AD dementia (N = 43 (37.4%)). AD with mixed pathology included AD with cerebral amyloid angiopathy (CAA), dementia with Lewy bodies (DLB), vascular disease, or a mixture of several of these pathologies. Non-AD dementia included TDP-43, α-syn, tau, or other non-AD pathologies (Table 2).

### 3.3 Comparison of p-tau217 plasma assays to CSF amyloid and tau biomarkers

P-tau217 concentrations of both plasma assays were significantly increased in the amyloid and tau positive (A+T+) group compared to the A-T-group. Amyloid positivity was determined by a CSF Aβ42/40 ratio of less than 0.073, including both positive (≤ 0.058) and likely positive (0.059 – 0.072) cases (Figure 1A, B). Tau positivity was determined by CSF p-tau181 levels > 50.2 pg/mL. This is consistent with other studies of amyloid and tau pathology for ALZpath and Fujirebio. ^30,31^ Encouragingly, both ALZpath and Fujirebio p-tau217 assays had very high correlation with both CSF amyloid (ALZpath: AUC 0.95; 95% CI 0.89 – 1.00, Fujirebio: AUC 0.94 (0.88 – 1.00)) and tau status (ALZpath: AUC 0.95 (0.90 – 1.00), Fujirebio: AUC 0.94 (0.88 – 1.00)) (Figure 1C, D). There was no significant difference in the performance of the two p-tau217 assays in predicting amyloid or tau abnormality compared to CSF.

**Figure 1:**
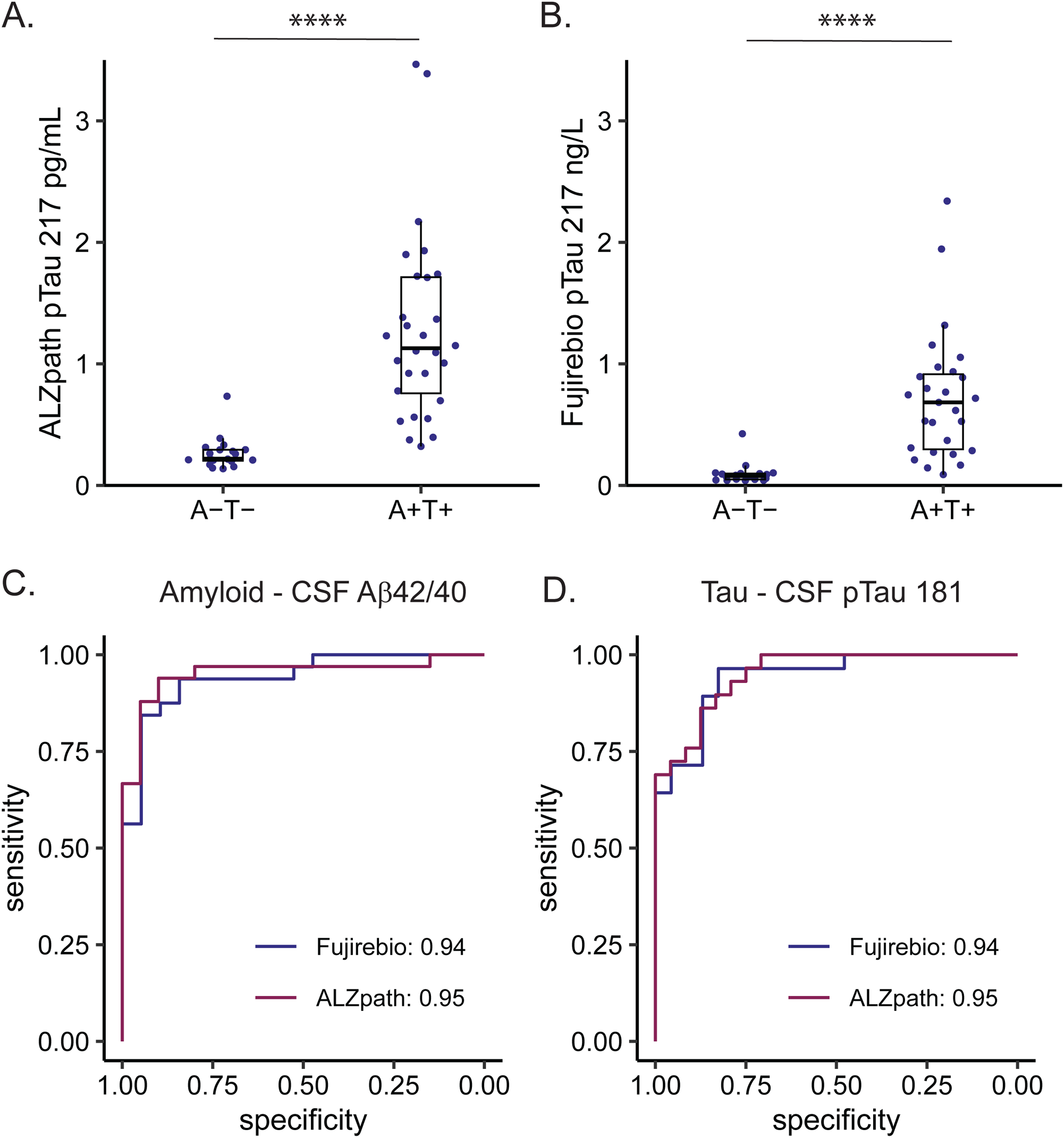
ALZpath and Fujirebio plasma p-tau217 assays have comparable clinical performance to CSF Aβ 42/40 and p-tau181 biomarkers. (**A, B**) p-tau217 concentration by AT profile for UBC CARD CSF cohort. Amyloid (A) was indexed by CSF Aβ 42/40 and tau (T) was indexed by CSF p-tau181. (**A**) ALZpath and (**B**) Fujirebio p-tau217 showed significantly increased concentrations in the A+T+ group, p-value < 0.0001. (**C**) ROC curve comparisons for plasma p-tau217 immuoassays compared to CSF Aβ42/40. (**D**) ROC curve comparisons for plasma p-tau217 immuoassays compared to CSF p-tau181.

Specifically, plasma p-tau217 correlates negatively with CSF Aβ 42/40 levels for both ALZpath (*r* = −0.72, p-value < 0.0001) and Fujirebio (*r* = −0.64, p-value < 0.001) (Supplemental figure 1). Similarly, plasma p-tau217 levels have a significant positive correlation with CSF p-tau181 levels, for ALZpath (*r* = 0.80, p-value < 0.001) and 0.42 for Fujirebio (r = 0.77, p-value < 0.0001) (Supplemental figure 1). A correlation between CSF and plasma AD biomarkers has been observed previously for plasma p-tau biomarkers.^32–36^

### 3.4 Diagnostic performance of plasma p-tau217 to identify autopsy-verified AD pathology from non-AD disorders

Using pathological diagnosis as the gold standard, the ALZpath plasma p-tau217 assay outperformed the Fujirebio assay for predicting autopsy-verified AD pathology, with a significantly higher AUC of 0.94 compared to 0.90 (p-value 0.023) (Figure 2A). For both platforms, plasma p-tau217 levels were significantly higher in autopsy-verified pure AD compared to AD with mixed disease pathology (p-value < 0.001 Figure 2B; p-value < 0.0001 Figure 2C) or non-AD disorders (p-value < 0.0001; Figure 2B & C). Moreover, the mixed AD cohort had significantly higher p-tau217 levels compared to non-AD disorders across both platforms (p-value < 0.0001 Figure 2B; p-value < 0.001 Figure 2C). However, the fold difference between the non-AD group and AD with mixed pathology is lower for the Fujirebio assay at 2.42 compared to 3.35-fold difference for ALZpath, whereas the fold difference between non-AD dementia and pure AD was higher for the Fujirebio assay at 6.93-fold compared to the ALZpath assay at 5.29-fold. Plasma p-tau217 is specific for AD pathology, including patients with a mixture of pathologies including CAA, DLB, vascular dementia, and a combination of three or more pathologies. This is a critical observation, because an NIA study found 54% of Alzheimer’s disease brain autopsies had coexisting pathology relating to other types of dementia in addition to AD pathology. ^37^ Patient with pure AD pathology or AD with CAA had the highest levels of p-tau217, followed by AD with DLB or vascular dementia. The lowest levels of p-tau 217 were observed in patients with a mixture of AD and at least two other pathologies associated with dementia. (Supplemental Figure 1).

**Figure 2:**
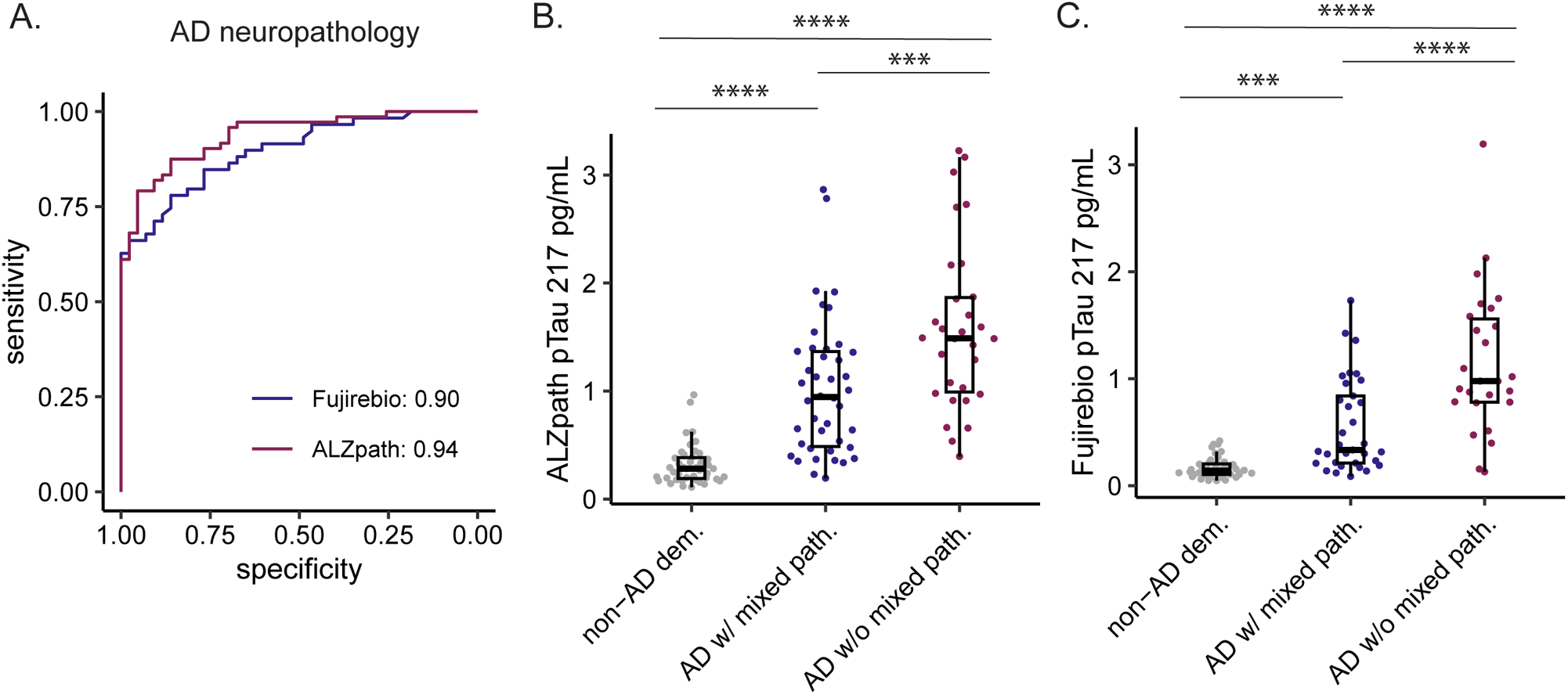
Comparison of ALZpath and Fujirebio plasma p-tau217 biomarker based on neuropathology. (**A**) ROC curve comparison for plasma p-tau217 immunoassays, Fujirebio AUC of 0.90 and ALZpath AUC of 0.94. (**B, C**) Pairwise comparison using Tuckey’s HSD test; *** p-value < 0.001 and **** p-value < 0.0001. (**B**) ALZpath p-tau217 levels in non-AD disease (N = 43), AD with mixed pathology (N = 42) and pure AD (N = 30). (**C**) Fujirebio p-tau217 levels in non-AD disease (N = 43), AD with mixed pathology (N = 33) and pure AD (N = 26).

### 3.5 Clinical decision points for plasma p-tau217 immunoassays

Separate decision points were set for the two p-tau217 tests based on the final diagnoses for the neuropathology cohort. ^38^Reference ranges for ALZpath p-tau217 have been previously established as < 0.40 ng/L and > 0.63 ng/L. ^30^ The specificity at the higher established threshold of 0.63 ng/L was 95.3%. Similar specificity (95.0%) was observed at 0.63 ng/L compared to amyloid pathology determined by CSF Aβ 42/40 ratio, suggesting this cut-off provides consistent results across cohorts and methods of determining amyloid pathology (Table 4). However, the lower established threshold of 0.40 ng/L showed < 90% sensitivity for amyloid pathology determined by both CSF Aβ 42/40 ratio and neuropathology. A lower threshold of 0.34 ng/L had a sensitivity of 95.8% for the neuropathology cohort and 93.4% for the CSF cohort, and a negative predictive value of 90.6% and 90.0%, respectively (Table 4). Within the intermediate zone (concentrations between 0.34 ng/L and 0.63 ng/L) 20.1% were within this range for the neuropathology confirmed cohort and 15.1% for the CSF cohort. For tau pathology determined by CSF p-tau181 the same reference ranges could be used, and there was 87.5% specificity and 88.0% positive predictive values at upper threshold 0.63 ng/L, with 93.1% sensitivity with 90.0% negative predictive value at lower threshold 0.34 ng/L.

**Table 4:** ALZpath plasma p-tau217 cut-offs.

Amyloid pathology - CSF Ab 42/40 - AUC 0.95
| Threshold | Spec. | Sens. | Acc. | PPV | NPV |
| --- | --- | --- | --- | --- | --- |
| 0.34 | 90.0% | 93.4% | 92.5% | 93.9% | 90.0% |
| 0.40 | 95.0% | 84.8% | 88.7% | 96.6% | 79.2% |
| 0.63 | 95.0% | 72.7% | 81.1% | 96.0% | 67.9% |

Tau pathology - CSF Ab p-tau181 - AUC 0.95
| Threshold | Spec. | Sens. | Acc. | PPV | NPV |
| --- | --- | --- | --- | --- | --- |
| 0.34 | 75.0% | 93.1% | 84.9% | 81.8% | 90.0% |
| 0.40 | 83.3% | 86.2% | 84.9% | 86.2% | 83.3% |
| 0.63 | 87.5% | 75.9% | 81.1% | 88.0% | 75.0% |

Amyloid neuropathology - AUC 0.94
| Threshold | Spec. | Sens. | Acc. | PPV | NPV |
| --- | --- | --- | --- | --- | --- |
| 0.34 | 67.4% | 95.8% | 85.2% | 83.1% | 90.6% |
| 0.40 | 76.7% | 87.5% | 83.5% | 86.3% | 78.6% |
| 0.63 | 95.3% | 79.2% | 85.2% | 96.6% | 73.2% |

Unique decision points were established for the Fujirebio p-tau217 assay. The lower threshold of 0.13 ng/L had 96.6% sensitivity and 90.9% negative predictive values, and the upper threshold of 0.37 ng/L had 93.0% specificity and 93.0% positive predictive values based on neuropathology (Table 5). However, only 87.5% sensitivity was observed at the lower threshold based on CSF Aβ 42/40 ratio, with similar specificity (94.7%) at the upper threshold (Table 5). These thresholds generate a larger intermediate zone compared to the ALZpath assay, with 32.2% of neuropathology cohort samples between 0.13 and 0.37, and 22.6% of the CSF cohort samples. A binary reference point is currently used by other clinical laboratories (0.18 ng/L) for this assay. For this decision point, we observed 67.4% specificity and 88.1% sensitivity for the neuropathology cohort and 94.7% specificity and 81.3% sensitivity for the CSF cohort. Similar sensitivity and specificity at 0.18 ng/L were observed for tau pathology (Table 5). These data suggest that the Fujirebio p-tau217 assay has less sensitivity than the ALZpath p-tau217 assay, and therefore a larger intermediate zone range would be required to achieve similar negative and positive predictive values.

**Table 5:** Fujirebio plasma p-tau217 cut-offs.

Amyloid pathology - CSF Ab 42/40 - AUC 0.94
| Threshold | Spec. | Sens. | Acc. | PPV | NPV |
| --- | --- | --- | --- | --- | --- |
| 0.13 | 84.2% | 87.5% | 86.3% | 90.3% | 80.0% |
| 0.18 | 94.7% | 81.3% | 86.3% | 96.3% | 75.0% |
| 0.37 | 94.7% | 59.4% | 72.5% | 95.0% | 58.1% |

Tau pathology - CSF Ab p-tau181 - AUC 0.94
| Threshold | Spec. | Sens. | Acc. | PPV | NPV |
| --- | --- | --- | --- | --- | --- |
| 0.13 | 82.6% | 96.4% | 90.2% | 87.1% | 95.0% |
| 0.18 | 87.0% | 85.7% | 86.3% | 88.9% | 83.3% |
| 0.37 | 95.7% | 67.9% | 80.4% | 95.0% | 71.0% |

Amyloid neuropathology - AUC 0.90
| Threshold | Spec. | Sens. | Acc. | PPV | NPV |
| --- | --- | --- | --- | --- | --- |
| 0.13 | 46.5% | 96.6% | 75.5% | 71.3% | 90.9% |
| 0.18 | 67.4% | 88.1% | 79.4% | 78.9% | 80.5% |
| 0.37 | 93.0% | 67.8% | 78.4% | 93.0% | 67.8% |

### 3.6 Plasma p-tau217 level and severity of AD by pathological staging

For both assays, the levels of p-tau217 increased according to Braak staging in the UBC-CARD neuropathology cohort. Plasma p-tau217 was significantly higher for individuals with Braak VI compared to individuals with Braak stage 0, I-II, III, IV, or V (Figure 3A, B). The ALZpath assay p-tau217 measurement increase was apparent by Braak stage IV and was nearly significant by Braak stage V (p-value = 0.07 Braak V to 0, and p-value = 0.09 Braak V to I & II). This increase was less pronounced with the Fujirebio p-tau217 assay (Figure 3B).

**Figure 3:**
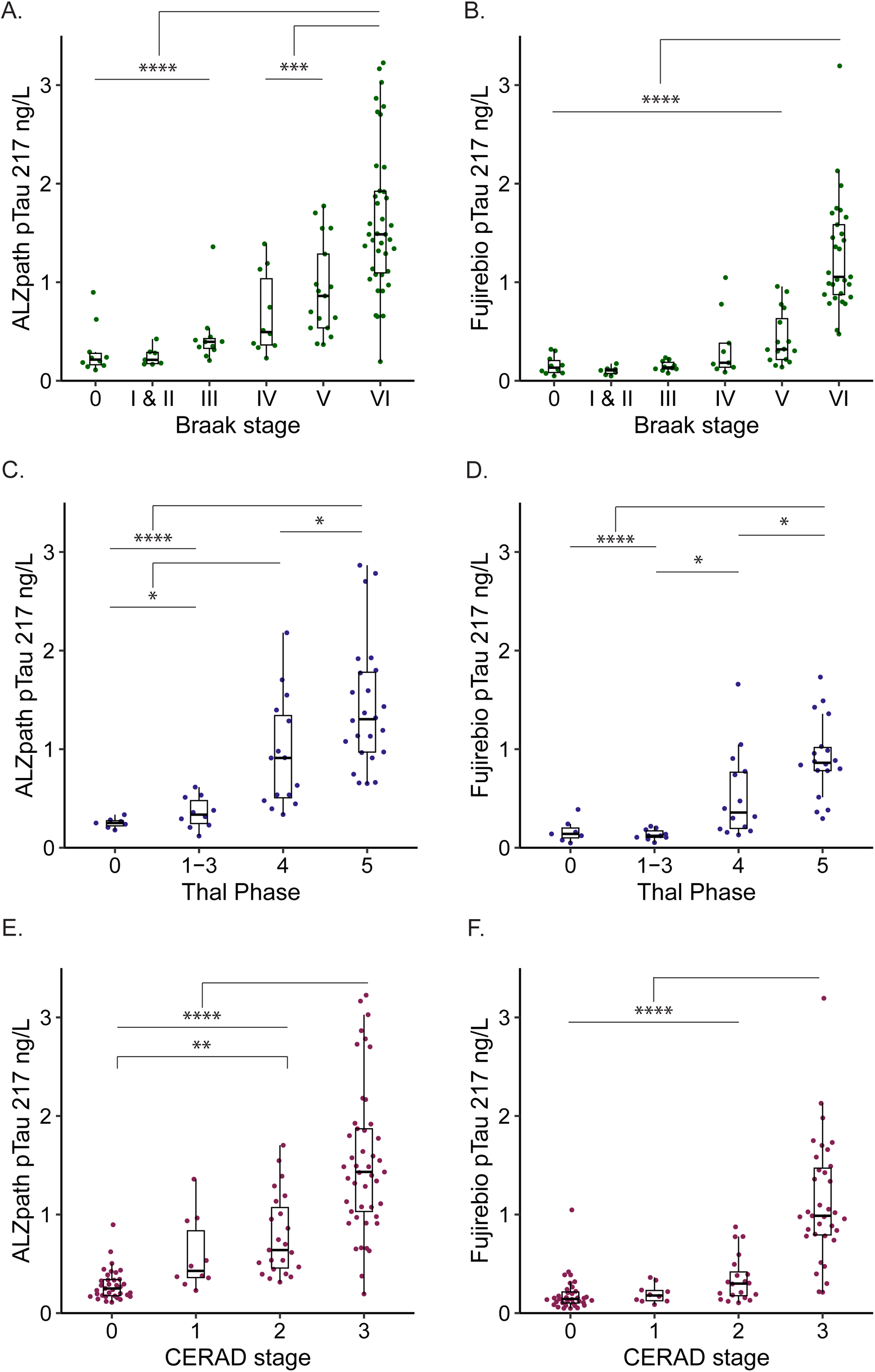
Neuropathologic evaluation of tau and amyloid-β scale compared to ALZpath and Fujirebio p-tau217 biomarkers. (**A, B**) Strong association with NFT burden based on Braak stages for both p-tau217 assays. (**A**) Visual inspection of the data suggests a biological effect in plasma p-tau217 at Braak stage III with the ALZpath assay and (**B**) stage IV with Fujirebio assay. (**C, D**) Diffuse Aβ plaque burden based on Thal phases. (**C**) Sequential increased at each Thal phase for ALZpath p-tau217 and (**D**) stage 4 and 5 for Fujirebio p-tau217. (**E, F**) Neuritic plaque location and density based on Consortium to Establish a Registry of Alzheimer’s Disease (CERAD) score. (**E**) Sequential increase at each CERAD stage observed for ALZpath p-tau217, (**F**) increase observed at CERAD stage 2 and 3 for Fujirebio p-tau217. Pairwise comparison using Tuckey’s HSD test; *p-value < 0.05, **p-value < 0.01, *** p-value < 0.001, and **** p-value < 0.0001.

Thal phase is a measure of diffuse beta amyloid throughout the brain. We found that p-tau217 was significantly higher in individuals at Thal phase 4 (p-value < 0.05) and 5 (p-value < 0.0001) compared to phase 3 and below (Figure 3C, D). The largest fold increase was observed at phase 4 with a 2.6-fold increase with the ALZpath assay and 4.1-fold increase with the Fujirebio assay. There was a significant difference between phase 4 and 5 with both the ALZpath and Fujirebio p-tau217 assays (p-value < 0.0001; ALZpath – 1.5x fold change; Fujirebio - 1.7x fold change). A slight, but non-significant, increase was observed between phase 0 and phase 1-3 for the ALZpath assay (p-value > 0.05; 1.4x fold change) (Figure 3C).

CERAD stage is determined based on the location and density of Aβ neurotic plaques. ^39^ ALZpath p-tau217 was increased in individuals with moderate plaque score at CERAD stage 2 (p-value < 0.01) and a substantial increase by CERAD stage 3 (p-value < 0.0001) (Figure 3E). This increase was only observed at CERAD stage 3 with the Fujirebio p-tau217 assay (p-value < 0.0001) and was not significant at earlier plaque stages (Figure 3F). These results show that the level of plasma p-tau217 for both ALZpath and Fujirebio increase according to AD disease staging, with increase observed at earlier disease staging with the ALZpath assay for both tau and amyloid pathology.

## 4. DISCUSSION

This study provides a head-to-head comparison of two immunoassays (ALZpath and Fujirebio) that have recently been made available for clinical use as laboratory-developed tests for plasma p-tau217. We assessed their analytical performance, as well as their correlations with CSF Aβ 42/40 and p-tau181 levels, and their diagnostic performance for AD and non-AD neurodegenerative diseases, confirmed by neuropathology. While both immunoassays exhibited excellent performance, the ALZpath assay demonstrated a slightly higher analytical and clinical performance with lower limits of detection and quantification, longer sample stability, and higher AUC compared to final neuropathology diagnosis. Our results demonstrate that both novel commercial p-tau217 immunoassays are able to identify amyloid and tau pathology, showing comparable performance to FDA-approved immunoassays for CSF Aβ42/40, used to indicate amyloid pathology, and RUO CSF p-tau181, used for tau pathology determination. ^40,41^

Current plasma p-tau217 biomarker assays exhibit great potential for AD diagnosis and staging.^15,18,30,42^ There are a few recent publications that clinically validated the plasma p-tau217 novel immunoassays including ALZpath. ^15,18,30,42^ In a recent published paper Benedet *et al.* compared the ALZpath p-tau217 assay with Janssen p-tau217+ assay in 294 individuals cross-sectionally, showing that both assays had excellent performance for AD diagnosis. ^43^ In another recent study, the Fujirebio Lumipulse and ALZpath SIMOA assays were compared by measuring plasma p-tau217 levels of 162 clinically diagnosed AD, 70 cases with other neurodegenerative diseases (NDDs) with CSF p-tau181 and 160 healthy controls. Both techniques showed a similar correlation with CSF p-tau181 levels and the same diagnostic accuracy for differentiating AD from NDDs.^22^ In a third recent study by Figdore *et al.* cut-offs for the ALZpath and Fujirebio p-tau217 immunoassays were assessed in clinically diagnosed MCI and mild AD cases determine by amyloid PET status. Using a one cut-point approach, neither assay achieved sensitivity and specificity ≥ 90%. However, with a two-cut point approach ≥ 90% sensitivity and specificity was achieved, and 39% of results were classified as indeterminate for the ALZpath assay and 22% for the Fujirebio. ^23^ Similar cut-off points were observed for the Fujirebio p-tau217 assay compared to our study, however, > 90% sensitivity was not observed at 0.18 pg/mL for either the CSF or neuropathology confirmed UBC-CARD cohorts. Comparing ALZpath p-tau217 cut-points between studies, the lower cut-point is similar to our study and the cut-point previously identified by Ashton *et al.*, however, the upper cut-point is significantly higher for Figdore *et al.* at 0.92 pg/mL compared to 0.63 pg/mL determine by our study and others. The reason for this discrepancy is unclear, but maybe due to differences in PET methodology in determining amyloid status or demographic difference in study cohorts. This suggests that further revaluation of the cut-points for both p-tau217 assay through prospective research studies are required before widespread clinical implementation.

It is noteworthy that all these studies evaluated the performance of plasma p-tau217 in comparison to amyloid PET and/or CSF. In our study, however, we compared the clinical performance of two recently developed plasma p-tau217 immunoassays against autopsy-confirmed cases, as well as CSF β42/40 and p-tau181 levels. Moreover, by using pathology-confirmed cases as a gold standard, the clinical validation of these two novel immunoassays showed that the sensitivity of the ALZpath assay in the diagnosis of AD is higher than that of the Fujirebio assay at the lower reference value for both assays. This difference in sensitivity may be due to a number of reasons, including the different antibodies used or platform methodology. Due to the lower sensitivity of the Fujirebio assay, there is greater overlap between the AD with mixed pathology and non-AD disease subgroups compared to the ALZpath assay. We speculate that patients who have additional neurodegenerative disease pathologies may require less severe amyloid and tau pathology to exhibit dementia symptoms and were therefore lower a p-tau217 levels. With the ALZpath assay, the levels of p-tau217 show an increase as early as Braak stage III & IV, and a stepwise increase at each CERAD stage. The Fujirebio assay has a modest increase at Braak stage IV, and no significant increase until CERAD stage 3. These results together suggest that the higher sensitivity of the ALZpath assay is able to detect amyloid and tau pathology at earlier disease staging compared to the Fujirebio assay.

However, the Fujirebio assay exhibits higher specificity for tau pathology at the upper reference value. This could be beneficial in primary care settings, where the rate of positivity is expected to be lower compared to specialized memory clinics. While plasma p-tau217 has shown promising discriminative accuracy for AD and is a strong candidate for implementation as a clinically diagnostic biomarker, this head-to-head study on various pathologically diagnosed neurodegenerative cases provides evidence that distinct p-tau217 assays may be necessary for screening, differential diagnosis, and disease monitoring in AD.

A limitation of our study is the retrospective nature of the design. Prospective studies with serial measurements are required to understand when p-tau217 levels start to deviate from the normal level when AD pathology develops. While plasma p-tau217 shows promise as a blood biomarker for AD diagnosis in real-world situations, it is crucial to investigate how pre-analysis variations and challenges in maintaining consistency can influence its reliability. An additional limitation of this study is the gap in time between sample collection and post-mortem brain autopsy, which can be up to several years.

In conclusion, this study, along with previous research, suggests the use of plasma p-tau217 as a diagnostic tool to distinguish AD pathology. We conducted a head-to-head comparison of two commercially available laboratory developed p-tau217 assays. Both assays demonstrate excellent analytical performance, and effectively differentiated AD pathology from non-AD neurodegenerative diseases. Overall, a 3-range approach can be used with the ALZpath p-tau217 assay while maintaining an intermediate zone of ≤ 20%, with follow up confirmation for the intermediate zone cases by PET or CSF testing. A 3-range approach for the Fujirebio p-tau217 assay results in an intermediate zone of > 20% to maintain the sample level of sensitivity and specificity. Further studies in real world memory clinics using these cut-off points are necessary to fully evaluate the effectiveness of plasma p-tau217 in routine clinical practice.

## Data Availability

All data produced in the present study are available upon reasonable request to the authors

## ACKNOWLEDGMENTS

The authors wish to thank the UBC-CARD study participants who took part in this research. We would like to thank Andreas Jeromin for his critical insight into the ALZpath p-tau217 assay, as well as Nathalie Le Bastard and Manu Vandijck for their critical insight into Fujirebio p-tau217 assay. We acknowledge the support of Neurocode USA Inc. for providing resources essential to the completion of this research.

## CONSENT STATEMENT

Ethical approval was sought from the Research Ethical Board (REB) at the University of British Columbia. Informed consent for the collection of relevant clinical information and the assessment of test results were obtained from the patients or their legal next of kin.

